# The roles of lifestyle factors and genetic risk in the association between night shift work and cholelithiasis: a prospective cohort study

**DOI:** 10.1101/2024.09.06.24313160

**Authors:** Wangping He, Ningning Mi, Kecheng Jin, Boru Jin, Ruyang Zhong, Zhen Liu, Yanyan Lin, Ping Yue, Bin Xia, Qiangsheng He, Fang Gao Smith, Jie Zhen, Jinqiu Yuan, Wenbo Meng

## Abstract

**Background:** Night shift work has been linked to various adverse health outcomes, but its relationship with incident cholelithiasis remains unclear. This study aims to investigate the association between night shift work and the risk of cholelithiasis, assess the potential modifying effects of genetic susceptibility, and explore the mediating roles of lifestyle factors.

**Methods:** A total of 219,810 subjects who were either in paid employment or self-employed were included in the UK Biobank. Information on current and lifetime employment were collected. Genetic risk was quantified and stratified by a polygenic risk score (PRS) incorporating 13 known cholelithiasis-associated loci. We used Cox proportional hazard models to investigate associations between night shift work and risk of cholelithiasis. Lifestyle factors measured at baseline were explored as potential mediators.

**Results:** During a median follow-up of 13.76 years, 6450 incidents of cholelithiasis were documented. Compared with day workers, the hazard ratio (HR) and 95% confidence interval (CI) of cholelithiasis was 1.09 (1.01, 1.17) for individuals with rarely/some night shifts and 1.18 (1.04, 1.35) for those with usual/permanent night shifts. Among the 62,558 participants who had reports on lifetime experience of night shift work, those with a higher frequency of night shifts and a longer length of each night shift were associated with an increased risk of cholelithiasis. Notably, individuals with usual/permanent night shifts and high genetic risk exhibited the highest risk of cholelithiasis (HR: 1.48, 95% CI: 1.21, 1.81), with day workers at low genetic risk serving as the reference. Mediation analysis indicated that a substantial portion (24.6%) of the association was mediated by BMI, followed by unhealthy alcohol intake (4.5%) and sedentary time (1.8%).

**Conclusions:** Night shift work is associated with an increased risk of cholelithiasis, with this relationship being largely mediated by lifestyle factors. These findings suggest that reducing the frequency and duration of night shifts may help mitigate the incidence of cholelithiasis among night shift workers, particularly for those with heightened genetic susceptibility.

## Introduction

Cholelithiasis is characterized by the presence of gallstones in the gallbladder or bile duct and affects 10 – 20% of the global adult population. In the United States, approximately 800,000 cholecystectomies are performed annually for this condition, contributing to an estimated healthcare cost of nearly 6.0 billion dollars^1^. Given the rising prevalence of cholelithiasis, both primary and secondary prevention through the management of risk factors is essential^2^. While numerous risk factors, including obesity, smoking, dietary habits, and physical inactivity, have been associated with the development of cholelithiasis^3^, it is crucial to identify and control emerging risk factors early to reduce the healthcare burden associated with this disease.

With the increasing demands of modern society and the specialization of labor, shift work is becoming more prevalent among employees^4^. Shift work encompasses labor performed outside conventional daytime hours, including evening, night, rotating, or irregular shifts. It is estimated that 15-20% of the working population in Europe and the United States are engaged in jobs that require night shifts^4^. Despite the necessity of shift work, substantial evidence suggests that night shift work disrupts the body’s circadian rhythm and adversely affects metabolism and hormone secretion^5–7^.

Previous studies have linked night shift work to elevated risks of various health conditions, including cardiovascular disease, metabolic disorders, and certain cancers^6,8–10^, with the impact potentially intensifying with prolonged exposure to night shifts. While the mechanisms through which night shift work contributes to cardiovascular disease have been extensively studied^11^, there remains a significant knowledge gap concerning its association with cholelithiasis and the underlying pathways involved.

In addition to environmental and lifestyle factors, genetic predisposition also contributes to the development of cholelithiasis^12^. Large-scale genome-wide association studies (GWAS) have identified multiple independent genetic loci associated with cholelithiasis risk in European populations^13,14^. Since genetic susceptibility may modify the relationship between environmental and lifestyle factors and chronic disease, we also investigated the combined effects of genetic susceptibility and night shift work on the incidence of cholelithiasis.

To address these knowledge gaps, we utilized data from the UK Biobank to investigate the relationships between exposure to night shift work and cholelithiasis risk among 219,810 participants. Additionally, we integrated information on in-depth lifetime employment to examine the impact of the length and frequency of night shifts on cholelithiasis risk. For gene-environment interactions, we constructed a specific polygenic risk score (PRS) for cholelithiasis to investigate the combined effects of night shift work and genetic variations on the cholelithiasis risk. Considering that night shift work negatively impacts health through suboptimal health lifestyles^15^ and certain lifestyle factors are likely to play a mediating role, simply adjusting for lifestyle factors as confounders may lead to overadjustment bias^16^. Our study further explored the mediating role of lifestyle factors between night shift work and cholelithiasis.

## Materials and Methods

### Study population

The study population was drawn from the UK Biobank, a large-scale prospective cohort study that recruited over 500,000 individuals aged 37 to 73 years from 22 assessment centers across England, Scotland, and Wales between 2006 and 2010^17^. At baseline assessment, participants provided electronic consent and completed a touch-screen questionnaire along with a face-to-face interview. Trained staff collected various measurements, including height and weight. Participants also provided detailed information on their lifestyle, medical conditions, work hours, and demographic background. Health professionals inquired about medical history, health status, and medication use. Further details about the UK Biobank have been extensively documented in prior studies^18,19^.

For our analyses, we focused on participants who were in paid employment or self-employed (n□=□286,248). After excluding participants with a history of cholelithiasis or cholecystectomy at baseline and missing lifestyle information and covariates, 219,810 participants were included in the primary analysis. Among them, 165,282 subjects were included in the subsequent gene-environment interaction analysis. Out of those, 62,558 subjects have complete lifetime employment information from the follow-up in 2015 through online questionnaires (**Supplementary Figure S1**).

### Exposure assessment

In the UK Biobank, shift work is defined as a work schedule outside normal daytime working hours of 9:00 AM to 5:00 PM, which means working afternoons, evenings or nights or rotating through these shifts. Participants who indicated paid employment or self-employment were then asked “Does your work involve shift work?”. If yes, participants were further asked whether their main job involved night shifts, defined as “…a work schedule that involves working through the normal sleeping hours (e.g., from 12:00 a.m. to 6:00 a.m.).” Response options for both questions included “never/rarely,” “sometimes,” “usually,” or “always,” with additional options for “prefer not to answer” and “do not know”^20^. Based on this information, participants’ current night shift work status was categorized as “day workers” “shift, but rarely/some night shifts” and “usual/permanent night shifts”.

In assessing lifetime employment, subjects reported each job ever worked, the number of years in each job, and the number of night shifts per month for each job. The detailed assessment method has been described in previous studies^8,20^. Based on the above information, we calculated the average frequency (number/per month) of night shifts and average length (hours) of each night shift for each participant.

### Outcome ascertainment

The primary outcome of interest in our study was cholelithiasis. Incident cholelithiasis was identified through linkage to the Health and Social Care Information Centre (in England and Wales) and the National Health Service Central Register (in Scotland). Outcome events were determined based on the International Classification of Diseases (ICD)-10 codes, specifically coded as K80.

### Assessment of polygenic risk score (PRS) to cholelithiasis

Genotyping in the UK Biobank was performed on two arrays, UK BiLEVE and UK Biobank Axiom. Genotyping, quality control, and imputation procedures have been previously described in detail^21^. The polygenic risk score (PRS) was computed using the cholelithiasis-associated single nucleotide polymorphisms (SNPs) identified in our previous study^22^. Detailed information about 13 independent SNPs is provided in **Supplementary Table S1**. In short, the number of alleles (0, 1, or 2) for each individual was multiplied by the effect size of SNPs associated with cholelithiasis, and then summed to derive PRS for all individuals in the UK Biobank^23^. The PRS was formulated as: PRS□=□β_1_□×□SNP_1_□+□β_2_□×□SNP_2_□+□…□+□β_n_□×□SNP_n_ (β_n_ represents the effect size of the n-th SNP, and SNPn is the genotype value for the n-th SNP, indicating the number of risk alleles (0, 1, or 2) present in the individual), where a higher score indicates a larger genetic susceptibility to cholelithiasis onset. We then determined whether participants were at high or low genetic risk for cholelithiasis based on the median value of PRS.

### Assessment of covariates and lifestyle factors

Age, gender, race, UK Biobank assessment centers, index of multiple deprivation, education, health rating, long-standing illness, multivitamin use and mineral use were considered as potential covariates. Index of multiple deprivation (IMD) reflecting socioeconomic status and different research centers acquired directly from the UK Biobank. Other sociodemographic characteristics (age, gender, race, education) were collected through questionnaires or verbal interviews at baseline and health status (health rating, long-standing illness, multivitamin use and mineral use) was collected through the self-reported medical history or hospitalization records.

Lifestyle factors included status of body mass index (BMI), physical activity, smoking status, alcohol intake, sleep duration, physical activity, sedentary time, and dietary characteristics. Weight and height were measured at baseline during the initial assessment center visit. Body mass index (BMI) was calculated as weight divided by height squared (kg/m2) at the initial assessment center visit. Smoking status, alcohol intake, sleep duration was assessed by the touch-screen questionnaire. Physical activity was assessed using the validated short International Physical Activity Questionnaire (IPAQ)^24^, which measures time spent on walking, and moderate and vigorous activities, which were also assessed by the touchscreen questionnaire. We used TV time as a measure of sedentary time^25^. Dietary characteristics was assessed based on the intake of 5 dietary components (fruits, vegetables, fish, red meat, and processed meat) following the American Heart Association Guideline^26^. Healthy diet was defined as adherence 2 or more ideal diet components (**Supplementary Table S2**).

### Statistical analysis

Baseline characteristics were presented as mean (standard deviation, SD) for continuous variables, and as a number (percentage) for categorical variables. The person-years of follow-up were calculated from the assessment date for each participant until the first diagnosis of cholelithiasis, death, or the date of last follow-up (31 MAY 2022), whichever came first.

We employed Cox proportional hazards models were used to estimate the hazard ratios (HRs) and 95% confidence intervals (CIs) for the association between current night shift work, average frequency of night shifts and average length of each night shift and incident cholelithiasis. Schoenfeld tests for the proportional hazard assumption were conducted in the UK Biobank, and no evidence of violation of this assumption was observed.

Three models with increasing adjustment were utilized to control potential confounders. Model 1 was stratified by age, sex, and UK Biobank assessment centers. Model 2 was further adjusted for model 1 plus race, index of multiple deprivation and education level. In model 3, we further adjusted for health rating, long-standing illness, multivitamin use and mineral use.

Kaplan-Meier analysis was performed to show the cumulative incidence rate across the different PRS categories in all participants. To evaluate to what extent the association of night shift work with cholelithiasis could be modified by genetic risk, joint effects were conducted for individuals with high and low genetic risk of cholelithiasis.

To enhance our understanding of how night shift work contributes to cholelithiasis, we probed the potential mediation pathways. Lifestyle factors associated with night shift work and/or cholelithiasis were considered as potential mediators. These mediators were adjusted in the Cox models to examine whether, and to what extent, the HRs between night shift work and cholelithiasis were attenuated. Cause mediation analyses were based on counterfactual framework, which formally defines both direct and indirect effects, and were more robust to the various limitations of traditional adjustment-based mediation analysis^27^. Natural indirect effect (NIE), natural direct effect (NDE), and total effect (TE) were estimated by combining mediation and outcome models that adjusted for covariates in model 3.

The selected lifestyle factors were considered as potential mediators in a 3-step analysis. First, we estimated the association of night shift work with each lifestyle factor using the multivariate-adjusted linear or logistic regression models. Second, we used multivariate-adjusted Cox regression models to assess the associations of lifestyle factors significantly associated with night shift work with the risk of cholelithiasis (**Supplementary Table S3-4**). Third, we conducted mediating analyses based on lifestyle factors that were significantly associated with night work shift and cholelithiasis.

To examine the robustness of the results, several sensitivity analyses were performed. Firstly, we excluded incident cases that occurred within the first year. which could reduce the possibility of reverse causation, enhance temporal considerations, and allow a time window for the development of cholelithiasis. We further adjusted chronotype category in the models. All statistical analyses were performed using R software (version 4.3.2) and Python (version 3.7.13), and a two-sided p-value < 0.05 was considered statistically significant.

## Results

**Table 1** presents the baseline characteristics of the study population according to current night shift status. In general, among 219,810 participants, night shift workers were more likely to be younger, male, nonwhite, less educated and more physically active and to have higher BMI, less healthy lifestyles, lower socioeconomic status and worse healthy status. The baseline characteristics of participants according to average monthly frequency of night shifts and average length of each night shift are presented in **Supplementary Table S5-6**. The participants possessing lifetime employment information exhibited comparable characteristics.

**Table 1.** Basic characteristics of participants by current work schedule.

| Characteristics | Day workers<br>(N=184817) | Shift, but rarely/some night shifts<br>(N=27518) | Usual/always night shifts<br>(N=7475) | Overall<br>(N=219810) |
| --- | --- | --- | --- | --- |
| **Mean (SD) Age, years** | 52.7 (7.09) | 51.7 (7.00) | 51.0 (6.77) | 52.5 (7.08) |
| **Gender, N (%)** |  |  |  |  |
| Female | 94079 (50.9) | 12180 (44.3) | 2597 (34.7) | 108856 (49.5) |
| Male | 90738 (49.1) | 15338 (55.7) | 4878 (65.3) | 110954 (50.5) |
| **Mean (SD) Index of multiple deprivation** | 15.6 (12.5) | 19.7 (14.9) | 21.8 (15.7) | 16.3 (13.0) |
| **Mean (SD) body mass index, kg/m<sup>2</sup>** | 26.9 (4.52) | 27.7 (4.80) | 28.3 (4.78) | 27.1 (4.58) |
| **Health rating, N (%)** |  |  |  |  |
| Excellent | 38871 (21.0) | 4337 (15.8) | 1113 (14.9) | 44321 (20.2) |
| Good | 111484 (60.3) | 16331 (59.3) | 4324 (57.8) | 132139 (60.1) |
| Poor | 3438 (1.9) | 760 (2.8) | 242 (3.2) | 4440 (2.0) |
| **Long-standing illness, N (%)** | 43420 (23.5) | 7250 (26.3) | 1951 (26.1) | 52621 (23.9) |
| **Education level, N (%)** |  |  |  |  |
| A levels/AS levels or equivalent | 24058 (13.0) | 3391 (12.3) | 841 (11.3) | 28290 (12.9) |
| College or University degree | 81664 (44.2) | 7502 (27.3) | 1267 (16.9) | 90433 (41.1) |
| CSEs or equivalent | 10410 (5.6) | 2569 (9.3) | 925 (12.4) | 13904 (6.3) |
| NVQ or HND or HNC or equivalent | 10956 (5.9) | 2578 (9.4) | 827 (11.1) | 14361 (6.5) |
| O levels/GCSEs or equivalent | 37304 (20.2) | 6712 (24.4) | 2074 (27.7) | 46090 (21.0) |
| Other | 6939 (3.8) | 1579 (5.7) | 545 (7.3) | 9063 (4.1) |
| **White, N (%)** | 175902 (95.2) | 24850 (90.3) | 6553 (87.7) | 207305 (94.3) |
| **Multivitamin use, N (%)** | 26105 (14.1) | 3924 (14.3) | 926 (12.4) | 30955 (14.1) |
| **Intake of mineral supplements, N (%)** | 41155 (22.3) | 6333 (23.0) | 1628 (21.8) | 49116 (22.3) |
| **Dietary characteristics, N (%)** |  |  |  |  |
| Unhealthy diet | 93734 (50.7) | 14471 (52.6) | 4190 (56.1) | 112395 (51.1) |
| Healthy diet | 91083 (49.3) | 13047 (47.4) | 3285 (43.9) | 107415 (48.9) |
| **Smoking status, N (%)** |  |  |  |  |
| Current | 17394 (9.4) | 3895 (14.2) | 1228 (16.4) | 22517 (10.2) |
| Never | 167423 (90.6) | 23623 (85.8) | 6247 (83.6) | 197293 (89.8) |
| **Alcohol intake, N (%)** |  |  |  |  |
| Healthy alcohol intake | 175203 (94.8%) | 25495 (92.6%) | 6794 (90.9%) | 207492 (94.4%) |
| Unhealthy alcohol intake | 9614 (5.2%) | 2023 (7.4%) | 681 (9.1%) | 12318 (5.6%) |
| **Physical activity, N (%)** |  |  |  |  |
| Healthy physical activity | 146714 (79.4) | 23411 (85.1) | 6513 (87.1) | 176638 (80.4) |
| Unhealthy physical activity | 38103 (20.6) | 4107 (14.9) | 962 (12.9) | 43172 (19.6) |
| **Sleep duration, N (%)** |  |  |  |  |
| Healthy sleep duration | 139745 (75.6) | 19007 (69.1) | 4530 (60.6) | 163282 (74.3) |
| Unhealthy sleep duration | 45072 (24.4) | 8511 (30.9) | 2945 (39.4) | 56528 (25.7) |
| **Watch TV, N (%)** |  |  |  |  |
| Healthy TV viewing time | 153273 (82.9) | 21508 (78.2) | 5426 (72.6) | 180207 (82.0) |
| Unhealthy TV viewing time | 31544 (17.1) | 6010 (21.8) | 2049 (27.4) | 39603 (18.0) |

During a median follow-up of 13.76 years, we documented 6450 incidents of cholelithiasis. We first tested the association of current night shift work status and cholelithiasis (**Table 2**). In age-, sex-, and UK Biobank assessment centers-stratified model 1, compared with day workers, the HR (95% CI) of cholelithiasis was 1.21 (1.13, 1.30) in shift workers with rarely/some night shifts and 1.36 (1.20, 1.55) in those with usual/permanent night shifts. In addition, this association was still significant after adjusting for race, index of multiple deprivation and education level in Model 2. Model 3 additionally adjusted for health rating, long-standing illness, multivitamin use and mineral use, and although estimates were further attenuated, results were overall comparable to model 2. In sensitivity analyses, after excluded cholelithiasis cases within the first year and further adjusted for chronotype category in the models (**Supplementary Table S7-8**), the results did not substantially differ from those observed in our aforementioned analyses.

**Table 2.** Current night shift work and cholelithiasis in the UK Biobank.

| Variables | HR (95% CI) by current work schedule |  |  | P for trend |
| --- | --- | --- | --- | --- |
|  | Day workers | Shift, but rarely/some night shifts | Usual/ permanent night shifts |  |
| Cases | 5292 | 899 | 259 |  |
| Person-years | 2485075.9 | 367718.5 | 100067.0 |  |
| Incidence rate* | 21.30 | 24.45 | 25.88 |  |
| Model 1 | 1.00 (ref) | 1.21 (1.13-1.30) | 1.36 (1.20-1.55) | <0.001 |
| Model 2 | 1.00 (ref) | 1.12 (1.04-1.21) | 1.22 (1.07-1.39) | <0.001 |
| Model 3 | 1.00 (ref) | 1.09 (1.01-1.17) | 1.18 (1.04-1.35) | 0.001 |

Abbreviations: ref= reference.
\*Per 10,000 person-years.
Model 1 stratified by age, sex and UK Biobank assessment centers; Model 2 further adjusted for race, index of multiple
deprivation and education level; Model 3 adjusted for terms in Model 2 and health rating, long-standing illness, multivitamin use
and mineral use.

Lifestyle factors were selected as potential mediators and these groups of mediators were adjusted in the Cox models to examine whether, and to what extent, the HRs between night shift work and cholelithiasis were attenuated (**Table 3**). Adjustments for the seven groups (dietary characteristics, smoking status, alcohol intake, physical activity, sleep duration, TV viewing, and BMI) of potential mediators are based on model 1. The strongest attenuation was seen following adjustment for BMI (the HR and 95% CI before adjustment: 1.09 [1.01,1.17] in□rarely/some night shifts and1.18 [1.04,1.35] in□rarely/some night shifts; after adjustment: 1.05 [0.98,1.13] in□rarely/some night shifts and1.12 [0.99,1.28] in□rarely/some night shifts).

**Table 3.** Association between current shift work and cholelithiasis by adjustment schemes.

| Variables | HR (95% CI) by current work schedule |  |  | P for trend |
| --- | --- | --- | --- | --- |
|  | Day workers | Shift, but rarely/some night shifts | Usual/ permanent night shifts |  |
| **Cases** | 5292 | 899 | 259 |  |
| **Person-years** | 2485075.9 | 367718.5 | 100067.0 |  |
| **Incidence rate<sup>e</sup>** | 21.30 | 24.45 | 25.88 |  |
| **Model 1** | 1.00 (ref) | 1.09 (1.01-1.17) | 1.18 (1.04-1.35) | 0.001 |
| **Model 2 (Model 1 + dietary characteristics)** | 1.00 (ref) | 1.09 (1.01-1.17) | 1.18 (1.04-1.35) | 0.001 |
| **Model 3 (Model 1 + smoking status)** | 1.00 (ref) | 1.08 (1.01-1.17) | 1.18 (1.04-1.34) | 0.002 |
| **Model 4 (Model 1 + alcohol intake)** | 1.00 (ref) | 1.09 (1.01-1.17) | 1.18 (1.04-1.34) | 0.001 |
| **Model 5 (Model 1 + physical activity)** | 1.00 (ref) | 1.10 (1.02-1.19) | 1.21 (1.06-1.37) | <0.001 |
| **Model 6 (Model 1 + sleep duration)** | 1.00 (ref) | 1.09 (1.01-1.17) | 1.18 (1.04-1.34) | 0.002 |
| **Model 7 (Model 1 + TV viewing<sup>†</sup>)** | 1.00 (ref) | 1.08 (1.01-1.17) | 1.17 (1.03-1.33) | 0.002 |
| **Model 8 (Model 1 + BMI)** | 1.00 (ref) | 1.05 (0.98-1.13) | 1.12 (0.99-1.28) | 0.032 |

Abbreviations: ref=reference, BMI= body mass index.
<sup>a</sup>Per 10,000 person-years.
<sup>†</sup>TV viewing was used to measure sedentary time.
Model 1 stratified by age, sex, and UK Biobank assessment centers and additionally adjusted for race, index of multiple
deprivation, education level, health rating, long-standing illness, multivitamin use and mineral use.

Mediation analyses were summarized in **Table 4**. Lifestyle factors associated with both night shift work and outcomes were selected for mediating analysis (**Supplementary Table S3-4**). The primary mediators identified were unhealthy alcohol intake (4.5%), unhealthy TV viewing or sedentary time (1.8%), and BMI (24.2%). Detailed mediation charts illustrating the natural indirect effect (NIE) and natural direct effect (NDE) of each lifestyle factor are provided in **Supplementary Figure S2.**

**Table 4.**
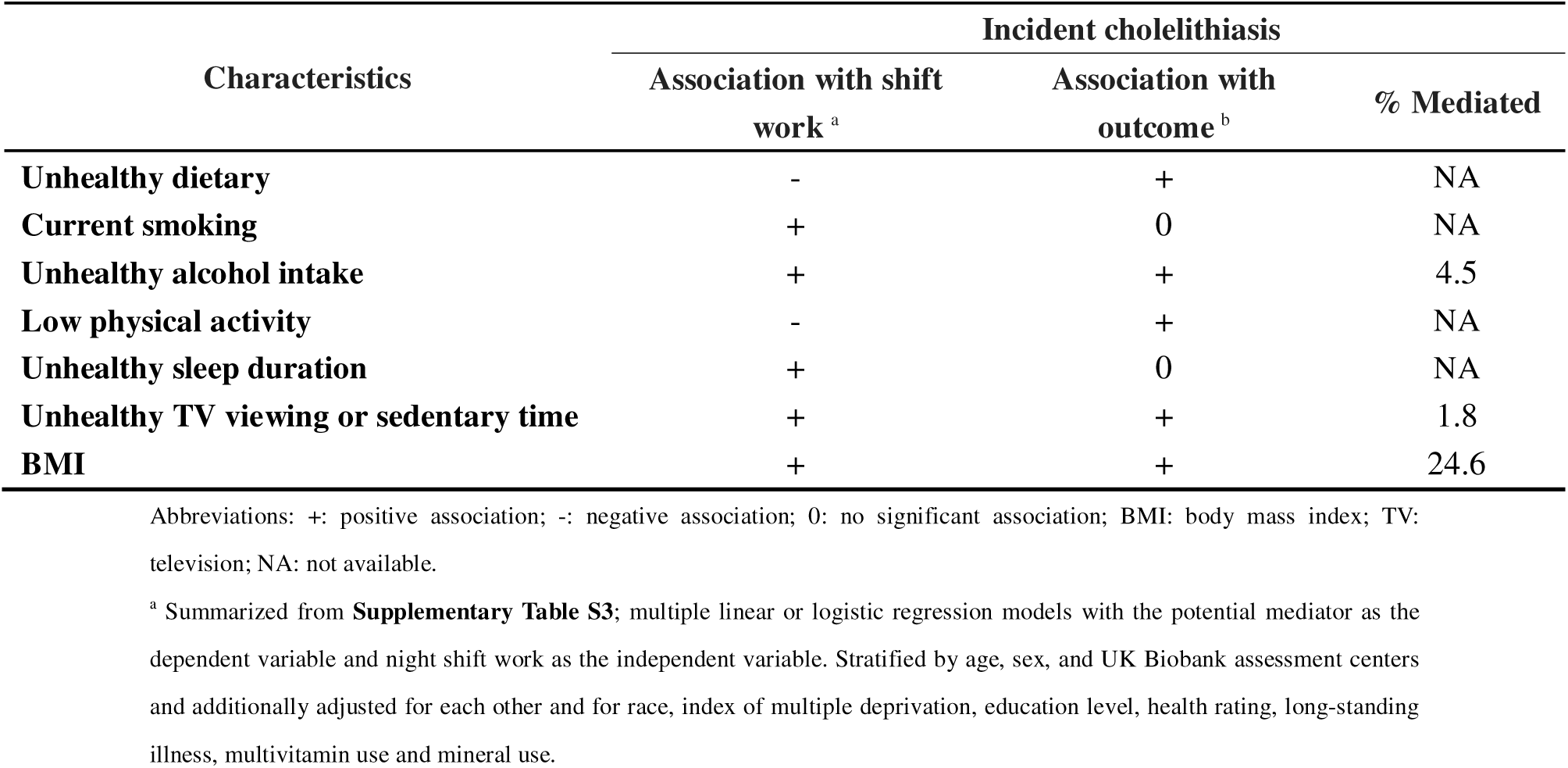
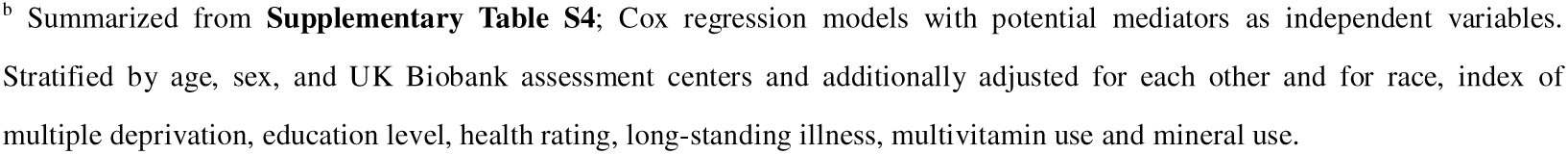
Summary of Mediation Analyses.

| Characteristics | Incident cholelithiasis |  |  |
| --- | --- | --- | --- |
|  | Association with shift work <sup>a</sup> | Association with outcome <sup>b</sup> | % Mediated |
| **Unhealthy dietary** | - | + | NA |
| **Current smoking** | + | 0 | NA |
| **Unhealthy alcohol intake** | + | + | 4.5 |
| **Low physical activity** | - | + | NA |
| **Unhealthy sleep duration** | + | 0 | NA |
| **Unhealthy TV viewing or sedentary time** | + | + | 1.8 |
| **BMI** | + | + | 24.6 |

Abbreviations: +: positive association; -: negative association; 0: no significant association; BMI: body mass index; TV:
television; NA: not available.
<sup>a</sup> Summarized from **Supplementary Table S3**; multiple linear or logistic regression models with the potential mediator as the
dependent variable and night shift work as the independent variable. Stratified by age, sex, and UK Biobank assessment centers
and additionally adjusted for each other and for race, index of multiple deprivation, education level, health rating, long-standing
illness, multivitamin use and mineral use.
<sup>b</sup> Summarized from **Supplementary Table S4**; Cox regression models with potential mediators as independent variables. Stratified by age, sex, and UK Biobank assessment centers and additionally adjusted for each other and for race, index of multiple deprivation, education level, health rating, long-standing illness, multivitamin use and mineral use.

We then examined the associations between lifetime night shift work (frequency and length) and the risk of cholelithiasis among 62,558 participants, with 1,578 incident cases. In fully adjusted models, we found that an increased frequency of night shifts was associated with a higher risk of cholelithiasis (**Table 5**). Compared with day workers, the HR (95% CI) of cholelithiasis was 1.18 (1.02, 1.37) in□< 8 night shifts/month and 1.26 (1.08, 1.46) in□≥8 night shifts/month. Additionally, we observed that night shift workers who undertook>12 h/shift had the highest cholelithiasis risk (HR 1.25 [95% CI 1.06, 1.48]) (**Table 6**).

**Table 5.** Average lifetime night shift frequency and risk of cholelithiasis.

| Variables | HR (95% CI) by average lifetime night shift frequency |  |  | P for trend |
| --- | --- | --- | --- | --- |
|  | None | <8/month | ≥8/month |  |
| Cases | 1122 | 232 | 224 |  |
| Person-years | 645107.98 | 111648.07 | 92601.68 |  |
| Incidence rate* | 17.39 | 20.78 | 24.19 |  |
| Model 1 | 1.00 (ref) | 1.26 (1.10-1.46) | 1.48 (1.28-1.71) | <0.001 |
| Model 2 | 1.00 (ref) | 1.20 (1.04-1.39) | 1.31 (1.13-1.53) | <0.001 |
| Model 3 | 1.00 (ref) | 1.18 (1.02-1.37) | 1.26 (1.08-1.46) | 0.001 |

Abbreviations: ref= reference.
\*Per 10,000 person-years.
Model 1 stratified for age, sex and UK Biobank assessment centers; Model 2 further adjusted for race, index of multiple deprivation and education level; Model 3 adjusted for terms in Model 2 and health rating, long-standing illness, multivitamin use and mineral use.

**Table 6.**
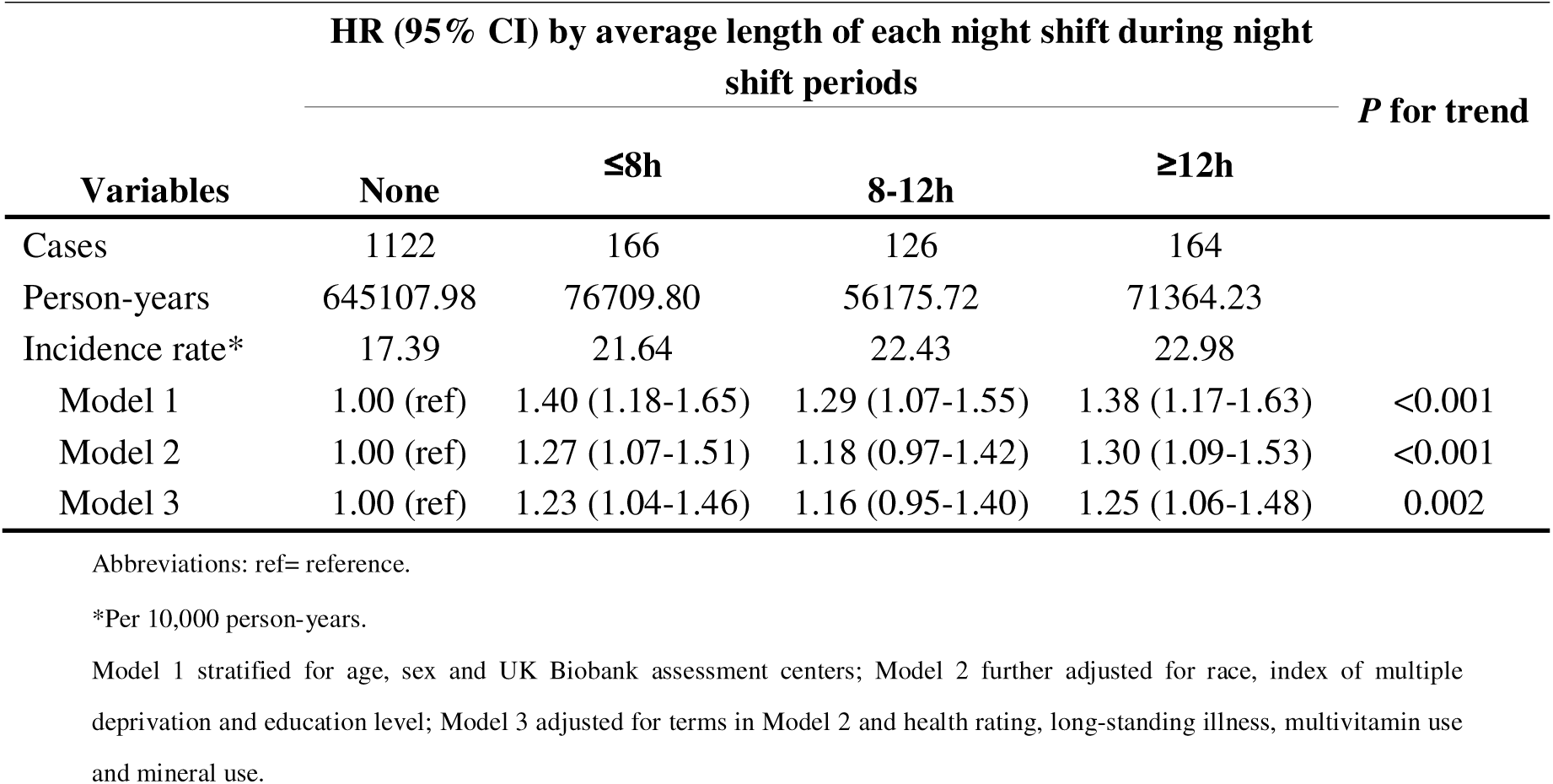
Average length of each night shift and risk of cholelithiasis.

The Kaplan-Meier curve showed that genetic risk was significantly associated with cholelithiasis (**Figure 1**). As expected, participants with high genetic risk had a broadly higher risk for cholelithiasis than those with a low genetic risk (HR 1.16, 95% CI [1.11,1.23]), suggesting that the PRS could effectively stratify risk for the cholelithiasis individuals. Furthermore, we observed significant joint effects between genetic risk and current work schedule on the cholelithiasis risk (**Figure 2**). Compared with day workers who had lower genetic risk, individuals with usual/permanent night shifts and high genetic risk had the highest risk of cholelithiasis (HR 1.48 [95% CI 1.21, 1.81]).

**Figure 1.**
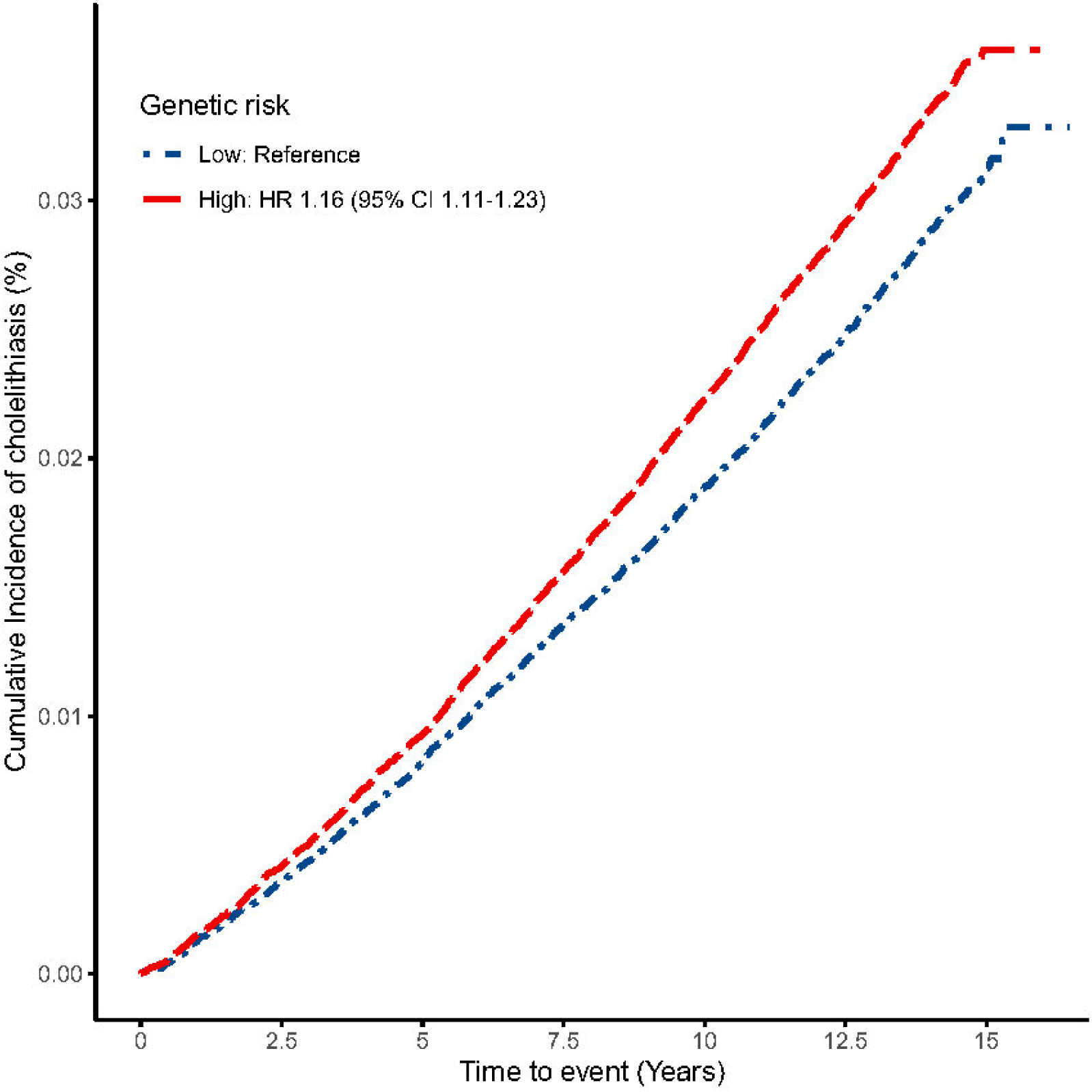
Kaplan-Meier curves of time to cumulative incidence of cholelithiasis by night shift work.

**Figure 2.**
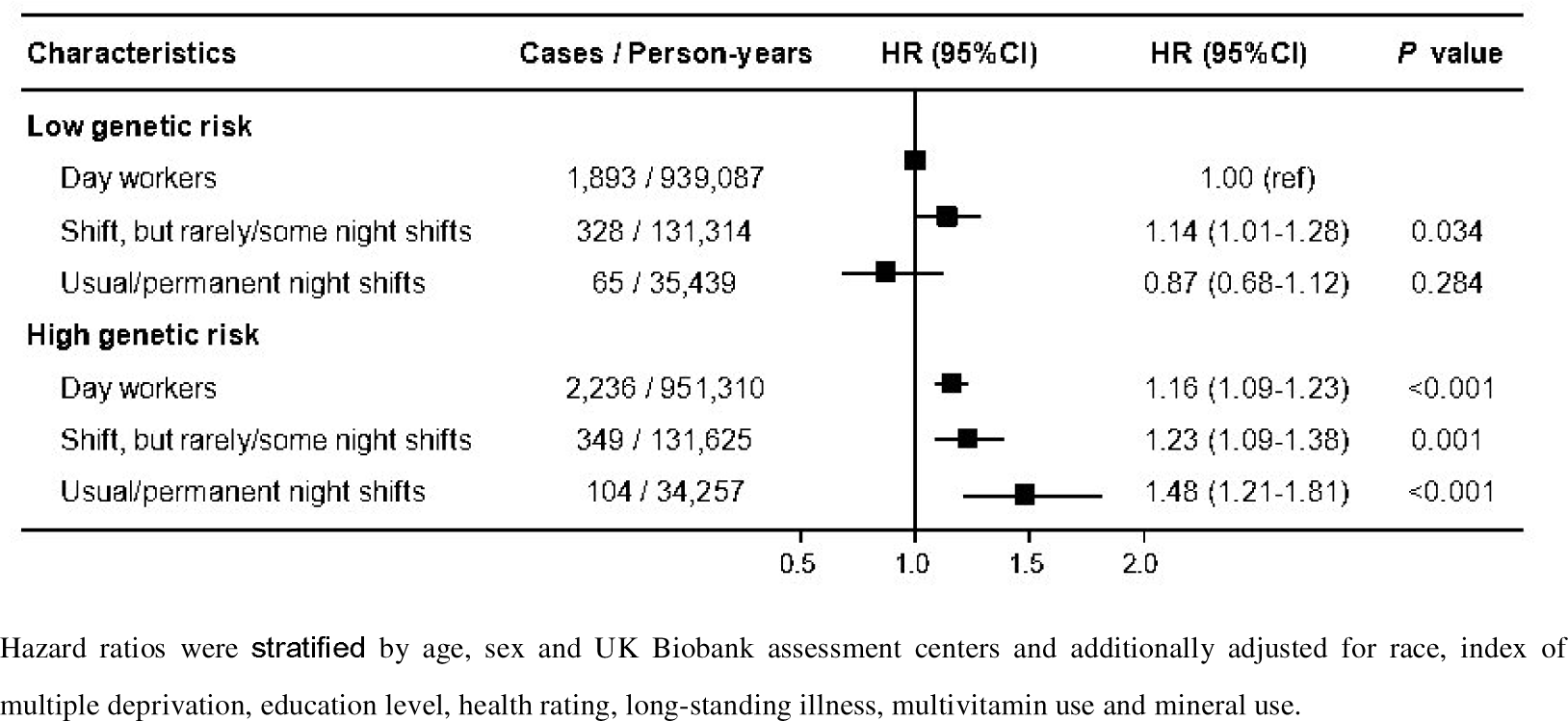
Joint effects of genetic risk with current work schedule on cholelithiasis risk.

## Discussion

In this study, we identified a significant association between night shift work and an increased risk of cholelithiasis. The risk showed a progressive increase from day work to irregular night shifts and regular night shifts. Joint analysis revealed that participants with usual/permanent night shift work and high genetic risk had the highest risk of incident cholelithiasis compared to those with day workers who had lower genetic risk. Additionally, longer length of each night shift and greater frequency of night shifts were related to a higher cholelithiasis risk. From a lifestyle perspective, BMI, alcohol intake, and sedentary time could partially mediate the association between night shift work and cholelithiasis, with the most notable mediating effect by BMI. These findings shed light on potential targets for interventions to reduce the subsequent risk of cholelithiasis in population who are at higher genetic risk.

To the best of our knowledge, this is the first prospective cohort study to investigate the relationships between night shift work and the risk of incident cholelithiasis. While previous research has extensively studied the association between night shift work and cardiovascular diseases^28,29^, cancers^30,31^, and metabolic diseases^6^, its adverse effects on the digestive system have been rarely mentioned. However, some limited studies have reported that associations between shift work and gastritis^32^, gastroesophageal reflux disease (GERD), irritable bowel syndrome(IBS), nonalcoholic fatty liver disease (NAFLD) ^33,34^, as well as higher odds of certain cancers^35,36^. To some extent, our results may be partly supported by those findings. For example, Sangyoon Lee et al. proposed that individuals working night shifts experienced more gastrointestinal symptom. They further found that those workers had a greater risk of gastritis compared to day workers^32^. Consistently, Hangkai Huang et al. found that the increased risk of NAFLD among individuals with night shift work was partly attributable to an increase in BMI^34^. Meanwhile, BMI is also recognized as an important risk factor for cholelithiasis^23^. In this study, we observed that the night shift was a significant and novel risk factor for cholelithiasis. These findings not only enhance our understanding of the digestive health impacts of night shifts but also provide crucial evidence for developing primary prevention strategies for cholelithiasis.

Furthermore, we noted that a higher frequency of night shifts and a longer length of each night shift and were important determinants of increased risk of cholelithiasis. Similarly, Eva et al. enrolled 78,586 women in the Nurses’ Health Study and demonstrated that the increasing years of night shifts was positively related to colorectal cancer risks^36^. Hangkai Huang et al. suggested that a longer duration, a higher frequency, more consecutive night shift work and longer hours per night shift were associated with higher risks of NAFLD^34^. All these findings provide evidence on detrimental relationships between night shift work and health and suggest that reducing the frequency and length of night shift work might be an effective measure to prevent digestive diseases in night shift workers.

The potential mechanism linking night shift work and cholelithiasis remains unclear, but there are several possible explanations. First, it is well known that circadian disruption, light exposure at night, and sleep deprivation are characteristic features of night shift work^10^. Evidence from animal studies has suggested that disordered circadian rhythm and abnormal cholesterol metabolism in mice promote gallstone formation^37^. Additionally, unhealthy sleep traits may increase the risk of gallbladder disease^38^. Exposure to light at night can lead to suppression of pineal hormone melatonin and increase in proinflammatory reactive oxygen species^10^. Meanwhile, inflammatory changes in gall bladder mucosa and reduced gall bladder motility are considered as risk factors for cholelithiasis^39^. As a result, Sreedevi et al. suggest that melatonin could be a viable treatment for treat gallstones, targeting reactive oxygen species and the hypomotility of the gall bladder^40^. Second, the expression of genes that encode key enzymes in hepatic bile acid and cholesterol metabolism, such as *Hmgcr* and *Cyp7a1*, is regulated by circadian rhythm-related transcription factors^41^. Therefore, the disturbance of circadian rhythm caused by night work can affect cholesterol metabolism and ultimately lead to the occurrence of cholelithiasis. In addition, the composition of the gut microbiota also shows circadian fluctuations^42^, which when disturbed by night work may contribute to the development of cholelithiasis. These findings suggest that the disturbance of intestinal microbiota among night shift workers may be one of the reasons for the increased incidence of cholelithiasis. Finally, the circadian disruption can disturb diurnal rhythms in vagal afferents^43^. The hepatic branch of the vagus nerve plays a critical role in biliary function. Damage to the hepatic branch can lead to impaired gallbladder contraction, the accumulation of bile salts, cholestasis, and eventually the formation of gallstones^44^. Therefore, the high incidence of cholelithiasis among night shift workers may be caused by abnormal vagal afferent rhythm.

In our study, after adjusting for lifestyle factors, the association between night shift work and cholelithiasis was partially attenuated. Mediation analysis indicated that lifestyle factors such as unhealthy alcohol intake, increased BMI, and sedentary behavior partially explained the relationship between night shift work and cholelithiasis risk. These findings suggest that while promoting healthy lifestyles may mitigate some adverse effects of night shift work, additional protective measures are necessary.

We also investigated the interaction between night shift work and genetic predisposition on cholelithiasis risk. As expected, individuals with high genetic risk who engaged in regular or permanent night shift work had the highest risk of developing cholelithiasis. These results highlight the need for personalized prevention strategies and suggest that future primary prevention efforts for night shift workers should consider individual genetic susceptibility.

The strength of our study lies in its prospective design, large sample size, long-term follow-up, and detailed current shift work information. Most importantly, this is the first cohort study to investigate relationships between night shift work and cholelithiasis risk, while also assessing the contribution of genetic factors in these relationships. This enabled us to precisely determine the effects of night shifts on groups with varying levels of susceptibility. Additionally, a series of sensitivity analyses were conducted to show the robustness of the findings. In addition, we have further clarified the roles of multiple lifestyle factors, helping to guide and improve strategy development and practice in health areas related to shift work.

Some limitations also remained in our study. First, as an observational study, it cannot establish causality between night shift work and cholelithiasis. Second, the exposure data was obtained from questionnaires which included whether participants were involved in night shift work and the frequency and length of shift work. This method may not be as accurate as measuring exposures using laboratory parameters. Third, the assessments of current and lifetime employment information in this study were based on baseline levels rather than being dynamic which may not accurately represent long-term status. In addition, despite careful adjustment for major confounders, bias from unknown and unmeasured confounding factors may still exist. Lastly, the study predominantly included white participants, limiting generalizability to other ethnic groups such as Asians and Blacks. Thus, caution should be exercised when generalizing these findings to the broader population.

## Conclusions

In conclusion, night shift work, particularly night shift work with higher frequency and longer length, was significantly associated with an increased risk of cholelithiasis and maybe a novel risk factor for cholelithiasis. Our study also revealed that the risk of cholelithiasis associated with night shift work is further exacerbated by higher genetic risk. These findings highlight the importance of reducing the burden of cholelithiasis through intervention of potential mediators.

## Authors’ contributions

Wangping He: Conceptualization, Methodology, Writing-Original Draft. Ningning MI: Software, Data Curation, Visualization, Funding acquisition. Kecheng Jin: Writing-Original Draft. Ruyang Zhong: Writing-Original Draft. Boru Jin: Writing-Original Draft. Zhen Liu: Writing-Original Draft. Yanyan Lin: Writing-Review & Editing. Ping Yue: Writing-Review & Editing. Bin Xia: Formal analysis, Funding acquisition. Qiangsheng He: Software, Validation. Jinqiu Yuan: Formal analysis, Supervision, Project administration, Funding acquisition. Wenbo Meng: Supervision, Project administration, Funding acquisition.

## Ethics approval and consent to participate

Each participant provided a written informed consent before data collection. The UK Biobank obtained ethics approval from the National Research Ethics Committee (REC ID: 16/NW/0274).

## Consent for publication

Not applicable.

## Data Availability

UK Biobank is an open access resource, and the study website https://www.ukbiobank.ac.uk/ has information on available data and access procedures. Data sets used for the analysis will be made available under reasonable requests.

## Abbreviations

BMI: Body Mass Index
CI: Confidence interval
GWASs: genome-wide association studies
HR: Hazard ratio
ICD: International Classification of Diseases
IMD: Index of multiple deprivation
NDE: Natural direct effect (NDE)
NIE: Natural indirect effect
PRS: Polygenic risk score
SNPs: Single nucleotide polymorphisms
TE: Total effect

## Acknowledgements

We thank all participants and researchers from the UK Biobank who make the study possible.

