## Supplementary material for "The roles of lifestyle factors and genetic risk in the association between night shift work and cholelithiasis: a prospective cohort study"

**Supplementary Table S1.** Information on genetic variants associated with cholelithiasis.

**Supplementary Table S2.** Assessment of lifestyle factors.

**Supplementary Table S3.** Associations between night shift work and potential mediators.

**Supplementary Table S4.** Association between all potential mediators and cholelithiasis and mediation proportions.

**Supplementary Table S5.** Basic characteristics of participants by average lifetime frequency of night shifts worked across all reported jobs (n = 62558).

**Supplementary Table S6.** Basic characteristics of participants by average lifetime length of each night shift worked across all reported jobs (n = 62558).

**Supplementary Table S7.** Adjusted HRs (95% CIs) of the risk of incident cholelithiasis by current work schedule after excluding participants with cholelithiasis occurred within 1 year of follow-up.

**Supplementary Table S8.** Associations of current shift work schedule and the risk of incident cholelithiasis further adjusted for chronotype.

**Supplementary Figure S1.** Flowchart of the study.

**Supplementary Figure S2.** Mediation effects of lifestyle factors on the association of night shift work and cholelithiasis.

**Supplementary Table S1. Information on genetic variants associated with cholelithiasis.**

| **ID** | **Chr** | **Position** | **Effect allele** | **Other allele** | | **P value** | **EAF** | | **Effect weight** |
| --- | --- | --- | --- | --- | --- | --- | --- | --- | --- |
| rs62140201 | 2 | 42214198 | A | G | 2.97E-06 | | 0.017823 | | 0.138004 |
| rs77273414 | 2 | 44062784 | A | G | 3.61E-08 | | 0.017216 | | -0.17926 |
| rs4401454 | 4 | 95741658 | G | A | 8.52E-08 | | 0.194986 | | 0.053809 |
| rs35093039 | 5 | 24061216 | A | G | 2.54E-06 | | 0.166293 | | 0.050292 |
| rs78488360 | 7 | 52968812 | T | C | 2.2E-06 | | 0.020549 | | -0.13876 |
| rs10832491 | 11 | 15652089 | A | C | 2.44E-06 | | 0.077328 | | -0.07244 |
| rs11020842 | 11 | 94317202 | A | G | 3.65E-06 | | 0.03732 | | 0.097566 |
| rs4765873 | 12 | 2109251 | T | C | 2.4E-06 | | 0.091698 | | 0.064969 |
| rs41276914 | 15 | 90347338 | C | T | 6.88E-07 | | 0.074388 | | -0.07726 |
| rs4468717 | 18 | 3457606 | T | C | 2.61E-06 | | 0.138178 | | -0.056 |
| rs516246 | 19 | 49206172 | C | T | 1.51E-08 | | 0.625012 | | -0.04653 |
| rs2618566 | 20 | 17844684 | G | T | 1.84E-08 | | 0.32278 | | -0.04881 |
| rs1800961 | 20 | 43042364 | T | C | 6.74E-68 | | 0.045473 | | 0.313022 |

**Supplementary Table S2. Assessment of lifestyle factors.**

| **UK Biobank** | | | |
| --- | --- | --- | --- |
| **Lifestyle factor** | **Questionnaire** | **Healthy habit** | **Unhealthy habit** |
| Smoking status | "Do you smoke tobacco now?" and "In the past, how often have you smoked tobacco?" | past or never smoker | current |
| Alcohol intake | “About how often do you drink alcohol?” | current | past or never drinker |
| Physical activity | International Physical Activity Questionnaire short form – total time of moderate or vigorous-intensity physical activity | ≥150 minutes /week | ＜150 minutes/week |
| Sleep duration | “About how many hours sleep do you get in every 24 hours?” | ≥7 or ≤9h/day | <7 or >9h/day |
| BMI | BMI was calculated as weight divided by height squared (kg/m2). | Expressed as a continuous variable | |
| TV viewing / sedentary time | “In a typical day, how many hours do you spend watching TV?” | < 4 h/day | ≥ 4 h/day |
| Dietary characteristics (From Touchscreen questionnaire at baseline. Healthy diet was defined as meeting 2 or more healthy diet items) | | | |
| Fruit | "About how many of …. would you eat per day?” Separate questions for pieces of fresh and dried fruit. | ≥ 4.5 servings/day | < 4.5 servings/day |
| Vegetable intake | "About how many of …. would you eat per day?” Separate questions for tablespoons of salad or cooked/raw vegetables. | ≥ 4.5 servings/day | < 4.5 servings/day |
| Total fish intake | "How often do you eat oily fish? (e.g. sardines, salmon, mackerel, herring)" | ≥2 servings/week | < 2 servings/week |
| Red meat intake | "How often do you eat…?” Separate questions for Beef / lamb or mutton / pork (excluding processed meats such as ham or bacon). | ≥5 servings/week | ＜5 servings/week |
| Processed meat intake | "How often do you eat processed meats (such as bacon, ham, sausages, meat pies, kebabs, burgers, chicken nuggets)?" | ≤ 2 servings/week | ＜2 servings/week |

**Supplementary Table S3. Associations between night shift work and potential mediators.**

|  | **Adjusted β (95% CI)** | ***P* value** |
| --- | --- | --- |
| Unhealthy dietary* | −0.029 (−0.054, −0.004) | 0.02 |
| Current smoking* | 0.25 (0.21, 0.28) | <0.001 |
| Unhealthy alcohol intake* | 0.20 (0.15, 0.24) | <0.001 |
| Low physical activity* | −0.44 (−0.47, -0.41) | <0.001 |
| Unhealthy sleep duration* | 0.29 (0.26, 0.31) | <0.001 |
| Unhealthy TV viewing time/sedentary time* | 0.098 (0.068, 0.128) | <0.001 |
| BMI | 0.37 (0.32, 0.42) | <0.001 |

β: standardized regression coefficients; TV: television; BMI: body mass index;

*binary variables modelled by logistic regressions; the exponentiation of βs are odds ratios.

Stratified by age, sex, and UK Biobank assessment centers and additionally adjusted for each other and for race, index of multiple deprivation, education level, health rating, long-standing illness, multivitamin use and mineral use.

**Supplementary Table S4. Association between all potential mediators and cholelithiasis and mediation proportions.**

|  | **Incident cholelithiasis** | | |
| --- | --- | --- | --- |
|  | **Outcome regressed by potential mediator** | | **Mediation analysis** |
|  | **HR (95% CI)** | **P value** | **% mediated** |
| Unhealthy dietary* | 1.15 (1.09-1.21) | <0.001 | NA |
| Current smoking* | 1.07 (0.99-1.16) | 0.087 | NA |
| Unhealthy alcohol intake* | 1.21 (1.10-1.34) | <0.001 | 4.5 |
| Low physical activity* | 1.23 (1.16-1.30) | <0.001 | NA |
| Unhealthy sleep duration* | 1.04 (0.98-1.10) | 0.186 | NA |
| Unhealthy TV viewing time/sedentary time* | 1.24 (1.17-1.32) | <0.001 | 1.8 |
| BMI | 1.08 (1.07-1.08) | <0.001 | 24.6 |

HR: hazard ratio; CI: confidence interval; TV: television; BMI: body mass index;

*binary variables modelled by logistic regressions;

Stratified by age, sex, and UK Biobank assessment centers and additionally adjusted for each other and for race, index of multiple deprivation, education level, health rating, long-standing illness, multivitamin use and mineral use.

**Supplementary Table S5. Basic characteristics of participants by average lifetime frequency of night shifts worked across all reported jobs (n = 62558).**

| **Characteristics** | **Average lifetime night shift frequency** | | | |
| --- | --- | --- | --- | --- |
|  | **None(N=47476)** | **<8/month(N=8252)** | **≥8/month(N=6830)** | **Overall(N=62558)** |
| **Mean (SD) Age, years** | 52.9 (6.85) | 52.0 (6.79) | 52.5 (6.91) | 52.7 (6.86) |
| **Gender, N (%)** |  |  |  |  |
| Female | 26551 (55.9%) | 3806 (46.1%) | 2678 (39.2%) | 33035 (52.8%) |
| Male | 20925 (44.1%) | 4446 (53.9%) | 4152 (60.8%) | 29523 (47.2%) |
| **Mean (SD) Index of multiple deprivation** | 14.1 (11.2) | 15.3 (12.1) | 16.5 (12.9) | 14.6 (11.6) |
| **Mean (SD) body mass index, kg/m^2^** | 26.3 (4.37) | 27.1 (4.67) | 27.6 (4.65) | 26.6 (4.47) |
| **Health rate, N (%)** |  |  |  |  |
| Excellent | 12436 (26.2%) | 1937 (23.5%) | 1427 (20.9%) | 15800 (25.3%) |
| Good | 28360 (59.7%) | 4947 (59.9%) | 4084 (59.8%) | 37391 (59.8%) |
| Poor | 588 (1.2%) | 127 (1.5%) | 140 (2.0%) | 855 (1.4%) |
| **Long-standing illness, N (%)** | 10509 (22.1%) | 2118 (25.7%) | 1887 (27.6%) | 14514 (23.2%) |
| **Education level, N (%)** |  |  |  |  |
| A levels/AS levels or equivalent | 6505 (13.7%) | 1315 (15.9%) | 840 (12.3%) | 8660 (13.8%) |
| College or University degree | 26867 (56.6%) | 3723 (45.1%) | 2699 (39.5%) | 33289 (53.2%) |
| CSEs or equivalent | 1487 (3.1%) | 338 (4.1%) | 436 (6.4%) | 2261 (3.6%) |
| NVQ or HND or HNC or equivalent | 2005 (4.2%) | 457 (5.5%) | 526 (7.7%) | 2988 (4.8%) |
| O levels/GCSEs or equivalent | 7755 (16.3%) | 1616 (19.6%) | 1561 (22.9%) | 10932 (17.5%) |
| **White, N (%)** | 46354 (97.6%) | 8020 (97.2%) | 6606 (96.7%) | 60980 (97.5%) |
| **Multivitamin use, N (%)** | 6756 (14.2%) | 1113 (13.5%) | 856 (12.5%) | 8725 (13.9%) |
| **Intake of mineral supplements, N (%)** | 10632 (22.4%) | 1851 (22.4%) | 1475 (21.6%) | 13958 (22.3%) |
| **Dietary characteristics, N (%)** |  |  |  |  |
| Unhealthy diet | 23504 (49.5%) | 4210 (51.0%) | 3550 (52.0%) | 31264 (50.0%) |
| Healthy diet | 23972 (50.5%) | 4042 (49.0%) | 3280 (48.0%) | 31294 (50.0%) |
| **Smoking status, N (%)** |  |  |  |  |
| Current | 3153 (6.6%) | 720 (8.7%) | 694 (10.2%) | 4567 (7.3%) |
| Never | 44323 (93.4%) | 7532 (91.3%) | 6136 (89.8%) | 57991 (92.7%) |
| **Alcohol intake, N (%)** |  |  |  |  |
| Healthy alcohol intake | 45383 (95.6%) | 7850 (95.1%) | 6548 (95.9%) | 59781 (95.6%) |
| Unhealthy alcohol intake | 2093 (4.4%) | 402 (4.9%) | 282 (4.1%) | 2777 (4.4%) |
| **Physical activity, N (%)** |  |  |  |  |
| Healthy physical activity | 37532 (79.1%) | 6749 (81.8%) | 5635 (82.5%) | 49916 (79.8%) |
| Unhealthy physical activity | 9944 (20.9%) | 1503 (18.2%) | 1195 (17.5%) | 12642 (20.2%) |
| **Sleep duration, N (%)** |  |  |  |  |
| Healthy sleep duration | 36956 (77.8%) | 6225 (75.4%) | 4878 (71.4%) | 48059 (76.8%) |
| Unhealthy sleep duration | 10520 (22.2%) | 2027 (24.6%) | 1952 (28.6%) | 14499 (23.2%) |
| **Watch TV, N (%)** |  |  |  |  |
| Healthy TV viewing time | 41346 (87.1%) | 7012 (85.0%) | 5580 (81.7%) | 53938 (86.2%) |
| Unhealthy TV viewing time | 6130 (12.9%) | 1240 (15.0%) | 1250 (18.3%) | 8620 (13.8%) |

**Supplementary Table S6. Basic characteristics of participants by average lifetime length of each night shift worked across all reported jobs (n = 62558).**

| **Characteristics** | **Average lifetime length of each night shift** | | | | |
| --- | --- | --- | --- | --- | --- |
|  | **None** | **≤8 h** | **8–12 h** | **≥ 12 h** | Overall |
| **Mean (SD) Age, years** | 52.9 (6.85) | 53.0 (6.99) | 51.8 (6.71) | 51.8 (6.74) | 52.7 (6.86) |
| **Gender, N (%)** |  |  |  |  |  |
| Female | 26551 (55.9%) | 1684 (29.7%) | 2250 (54.3%) | 2550 (48.5%) | 33035 (52.8%) |
| Male | 20925 (44.1%) | 3992 (70.3%) | 1897 (45.7%) | 2709 (51.5%) | 29523 (47.2%) |
| **Mean (SD) Index of multiple deprivation** | 14.1 (11.2) | 16.3 (12.7) | 15.8 (12.3) | 15.4 (12.3) | 14.6 (11.6) |
| **Mean (SD) body mass index, kg/m^2^** | 26.3 (4.37) | 27.6 (4.51) | 27.2 (4.72) | 27.2 (4.77) | 26.6 (4.47) |
| **Health rate, N (%)** |  |  |  |  |  |
| Excellent | 12436 (26.2%) | 1183 (20.8%) | 894 (21.6%) | 1287 (24.5%) | 15800 (25.3%) |
| Good | 28360 (59.7%) | 3429 (60.4%) | 2558 (61.7%) | 3044 (57.9%) | 37391 (59.8%) |
| Poor | 588 (1.2%) | 112 (2.0%) | 64 (1.5%) | 91 (1.7%) | 855 (1.4%) |
| **Long-standing illness, N (%)** | 10509 (22.1%) | 1528 (26.9%) | 1079 (26.0%) | 1398 (26.6%) | 14514 (23.2%) |
| **Education level, N (%)** |  |  |  |  |  |
| A levels/AS levels or equivalent | 6505 (13.7%) | 855 (15.1%) | 677 (16.3%) | 623 (11.8%) | 8660 (13.8%) |
| College or University degree | 26867 (56.6%) | 2253 (39.7%) | 1625 (39.2%) | 2544 (48.4%) | 33289 (53.2%) |
| CSEs or equivalent | 1487 (3.1%) | 307 (5.4%) | 242 (5.8%) | 225 (4.3%) | 2261 (3.6%) |
| NVQ or HND or HNC or equivalent | 2005 (4.2%) | 430 (7.6%) | 262 (6.3%) | 291 (5.5%) | 2988 (4.8%) |
| O levels/GCSEs or equivalent | 7755 (16.3%) | 1294 (22.8%) | 866 (20.9%) | 1017 (19.3%) | 10932 (17.5%) |
| **White, N (%)** | 46132 (97.2%) | 5455 (96.1%) | 4004 (96.6%) | 5036 (95.8%) | 60627 (96.9%) |
| **Multivitamin use, N (%)** | 6756 (14.2%) | 768 (13.5%) | 535 (12.9%) | 666 (12.7%) | 8725 (13.9%) |
| **Intake of mineral supplements, N (%)** | 10632 (22.4%) | 1279 (22.5%) | 928 (22.4%) | 1119 (21.3%) | 13958 (22.3%) |
| **Dietary characteristics, N (%)** |  |  |  |  |  |
| Unhealthy diet | 23504 (49.5%) | 3007 (53.0%) | 2120 (51.1%) | 2633 (50.1%) | 31264 (50.0%) |
| Healthy diet | 23972 (50.5%) | 2669 (47.0%) | 2027 (48.9%) | 2626 (49.9%) | 31294 (50.0%) |
| **Smoking status, N (%)** |  |  |  |  |  |
| Current | 3153 (6.6%) | 548 (9.7%) | 370 (8.9%) | 496 (9.4%) | 4567 (7.3%) |
| Never | 44323 (93.4%) | 5128 (90.3%) | 3777 (91.1%) | 4763 (90.6%) | 57991 (92.7%) |
| **Alcohol intake, N (%)** |  |  |  |  |  |
| Healthy alcohol intake | 45383 (95.6%) | 5451 (96.0%) | 3947 (95.2%) | 5000 (95.1%) | 59781 (95.6%) |
| Unhealthy alcohol intake | 2093 (4.4%) | 225 (4.0%) | 200 (4.8%) | 259 (4.9%) | 2777 (4.4%) |
| **Physical activity, N (%)** |  |  |  |  |  |
| Healthy physical activity | 37532 (79.1%) | 4645 (81.8%) | 3431 (82.7%) | 4308 (81.9%) | 49916 (79.8%) |
| Unhealthy physical activity | 9944 (20.9%) | 1031 (18.2%) | 716 (17.3%) | 951 (18.1%) | 12642 (20.2%) |
| **Sleep duration, N (%)** |  |  |  |  |  |
| Healthy sleep duration | 36956 (77.8%) | 4142 (73.0%) | 3089 (74.5%) | 3872 (73.6%) | 48059 (76.8%) |
| Unhealthy sleep duration | 10520 (22.2%) | 1534 (27.0%) | 1058 (25.5%) | 1387 (26.4%) | 14499 (23.2%) |
| **Watch TV, N (%)** |  |  |  |  |  |
| Healthy TV viewing time | 41346 (87.1%) | 4679 (82.4%) | 3433 (82.8%) | 4480 (85.2%) | 53938 (86.2%) |
| Unhealthy TV viewing time | 6130 (12.9%) | 997 (17.6%) | 714 (17.2%) | 779 (14.8%) | 8620 (13.8%) |

**Supplementary Table S7. Adjusted HRs (95% CIs) of the risk of incident cholelithiasis by current work schedule after excluding participants with cholelithiasis occurred within 1 year of follow-up.**

| **Variables** | **HR (95% CI) by current work schedule** | | | ***P* for trend** |
| --- | --- | --- | --- | --- |
|  | **Day workers** | **Shift, but rarely/some night shifts** | **Usual/permanent night shifts** |  |
| Model 1 | 1.00 (ref) | 1.20 (1.11-1.29) | 1.38 (1.22-1.57) | <0.001 |
| Model 2 | 1.00 (ref) | 1.11 (1.03-1.20) | 1.24 (1.09-1.41) | <0.001 |
| Model 3 | 1.00 (ref) | 1.07 (1.00-1.16) | 1.20 (1.05-1.37) | 0.002 |

Abbreviations: ref= reference.

Model 1 stratified by age, sex and UK Biobank assessment centers; Model 2 further adjusted for race, index of multiple deprivation and education level; Model 3 adjusted for terms in Model 2 and health rating, long-standing illness, multivitamin use and mineral use.

**Supplementary Table S8. Associations of current shift work schedule and the risk of incident cholelithiasis further adjusted for chronotype.**

| **Variables** | **HR (95% CI) by current work schedule** | | | ***P* for trend** |
| --- | --- | --- | --- | --- |
|  | **Day workers** | **Shift, but rarely/some night shifts** | **Usual/ permanent night shifts** |  |
| Model 1 | 1.00 (ref) | 1.21 (1.13-1.30) | 1.36 (1.20-1.55) | <0.001 |
| Model 2 | 1.00 (ref) | 1.12 (1.04-1.21) | 1.22 (1.07-1.39) | <0.001 |
| Model 3 | 1.00 (ref) | 1.07 (0.99-1.16) | 1.15 (1.01-1.32) | 0.01 |

Abbreviations: ref= reference.

Model 1 stratified by chronotype, age, sex and UK Biobank assessment centers; Model 2 further adjusted for race, index of multiple deprivation and education level; Model 3 adjusted for terms in Model 2 and health rating, long-standing illness, multivitamin use and mineral use.

All modes further adjusted for chronotype category.

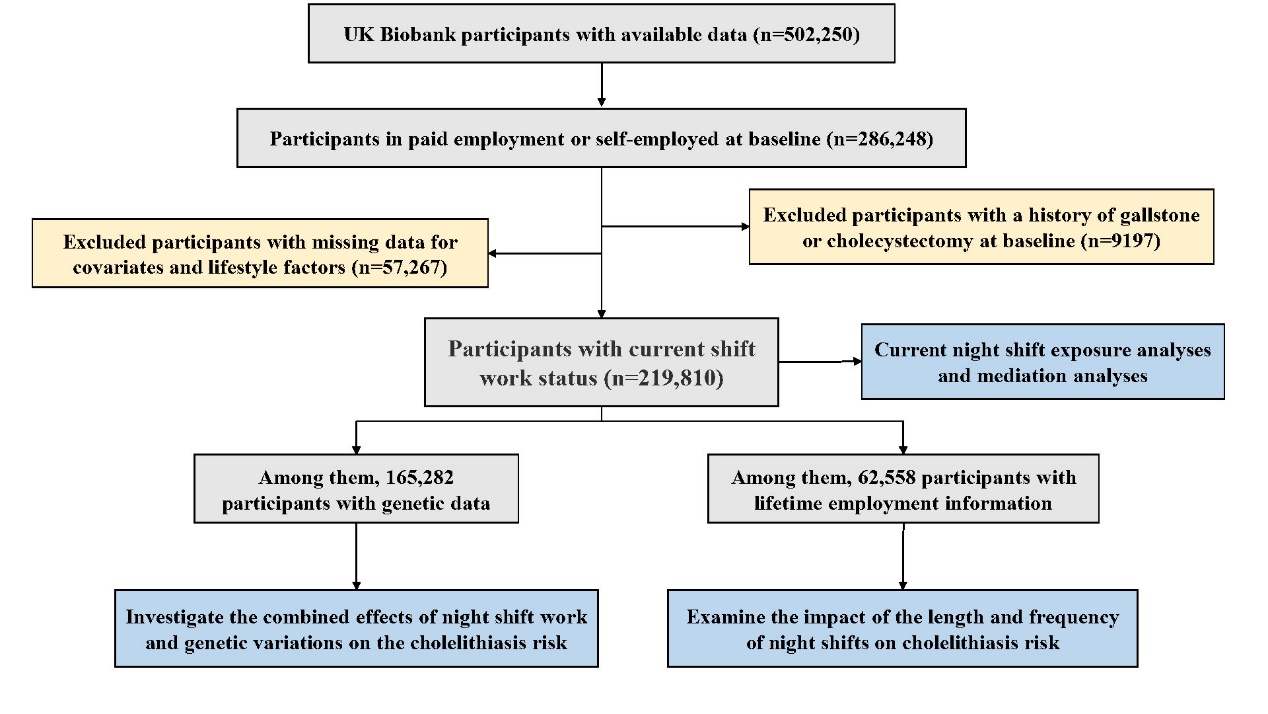

**Supplementary Figure S1. Flowchart of the study.**

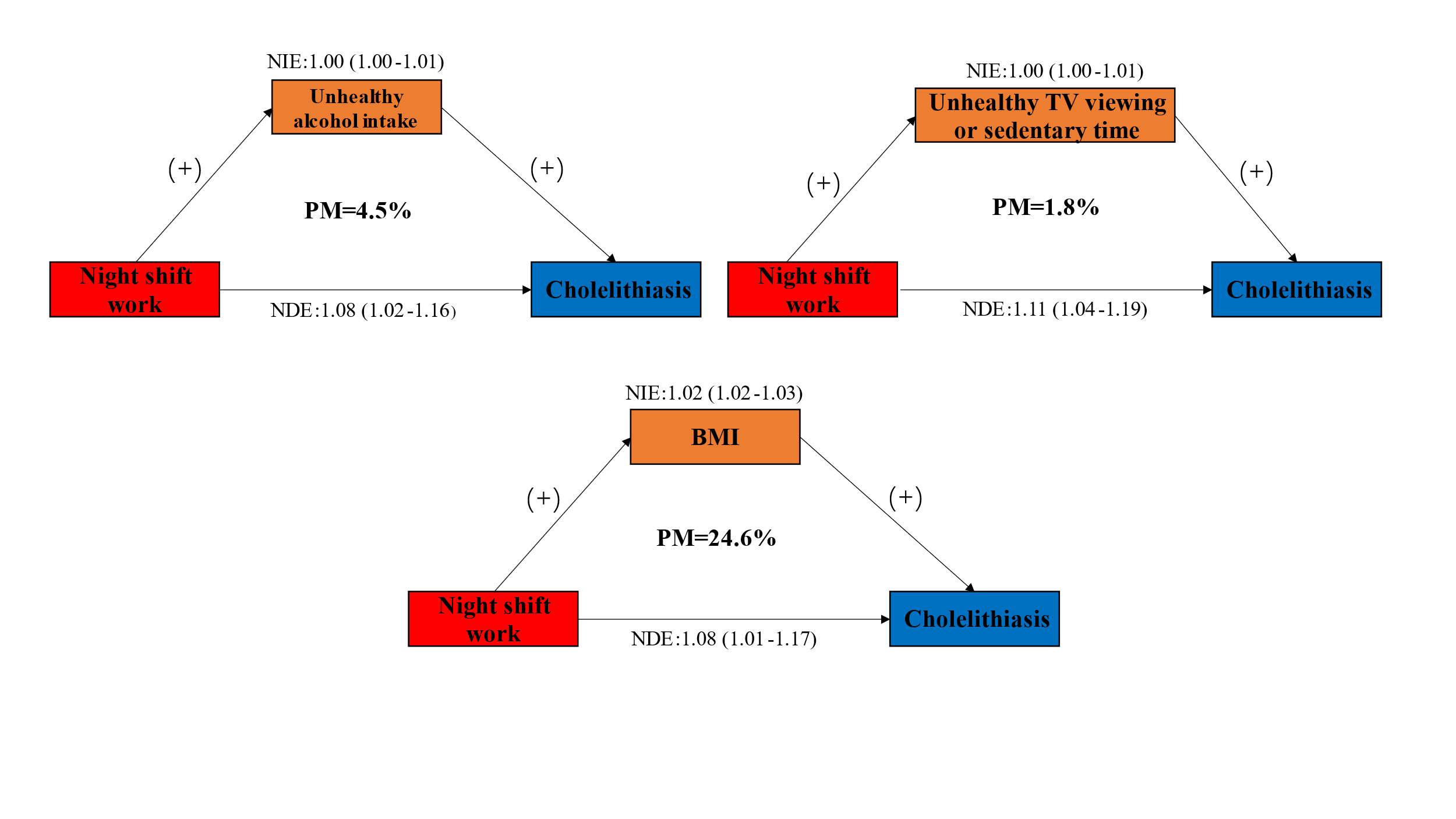

**Supplementary Figure S2. Mediation effects of lifestyle factors on the association of night shift work and cholelithiasis.**

Abbreviations: (+): indicates a positive association; PM: percent mediated; BMI: body mass index; NIE: natural indirect effect; NDE: natural direct effect.

Model stratified by age, sex, and UK Biobank assessment centers and additionally adjusted for race, index of multiple deprivation, education level, health rating, long-standing illness, multivitamin use and mineral use.
